# Improvements in patient-reported outcomes after neuromuscular electrical stimulation therapy for snoring and mild sleep apnea

**DOI:** 10.64898/2026.11.02.26367068

**Authors:** M.R. Crawford, A.L. Jones, S. Parthasarathy, C. Osborne, H.K. Walia, J. Ojile, A. Malhotra

**Affiliations:** Strathclyde Sleep Centre, Department of Psychological Sciences and Health, University of Strathclyde, Glasgow, UK; Department of Psychology, Swansea University, Swansea, UK; University of Arizona, Tucson, USA; Signifier Medical Technologies, London, UK; Baptist Health, Miami, USA; Clayton Sleep Institute, St. Louis, USA; UC San Diego Health, San Diego, USA

**Keywords:** Obstructive sleep apnea, neuromuscular electrical stimulation, patient reported outcomes

## Abstract

**Objectives:** Neuromuscular electrical stimulation (NMES) offers alternative options to patients with Obstructive Sleep Apnea (OSA) unable to tolerate first line treatment. Randomized controlled trial data suggest efficacy of NMES treatment in improving OSA outcomes, snoring and sleepiness; however, no study to our knowledge has examined the effects on a range of daytime patient reported outcomes in a large real-world sample.

**Methods:** This report presents clinical data from 472 patients completing NMES treatment. Bayesian linear mixed models were used to analyze pre-to-post treatment changes in mood (PHQ8), sleep disturbance (PROMIS-SD-4a), sleep related impairment (PROMIS-SRI-4a), and functioning (FOSQ), whilst controlling for key covariates. Posterior probabilities were calculated to estimate the probability of a change in symptoms (P(effect > 0)) and probability of exceeding a minimal clinically important difference (P(effect > MCID)).

**Results:** The posterior probabilities of an improvement in scores in sleep disturbance (mean change −6.1, 94% HDI: −3.8 to −8.5) and sleep-related impairment (mean change −10.0, 94% HDI: −5.4 to −14), depression (mean change −3.5, 94% HDI: −1.7 to −5.2), at posttreatment were greatest and varied between 99.8 to 100%. Probability of change in functioning was low (60.9% chance of improvement). These effects remained when controlling for age, gender, treatment adherence, mean intensity and variance in intensity. Probabilities for MCI change for mood, sleep disturbance and sleep-related impairment were high (86-91%).

**Conclusions:** The results from this report highlight the potential impact NMES therapy has on patient reported outcomes. These findings are in support of prior randomized placebo-controlled data and several observational cohort studies for NMES as treatment for mild OSA.

## Introduction

Obstructive sleep apnoea (OSA) is a sleep-related breathing disorder, characterized by repeated reduction (hypopnea) or cessation (apnoea) in airflow during sleep. Associated sleep disruption and persistent hypoxia may lead to negative health sequelae (Xie et al., 2017) increased risks of accidents (Garbarino et al., 2016; Luzzi et al., 2022), increased risk of Parkinson’s disease (Sun et al., 2020), poorer quality of life (Shirisha et al., 2025) and increased risk of depression (Edwards et al., 2020), which yields a major economic burden (Borsoi et al., 2022). While the gold-standard treatment Positive Airway Pressure eliminates most of the breathing events, adherence is suboptimal in many patients (Crew et al., 2021; Oliver et al., 2024; Rotenberg et al., 2016). While oral appliances are generally more easily tolerated than PAP therapy, effects on dental or occlusal structures remain a concern. Novel alternatives to PAP therapy and oral appliances have emerged, with a particular focus on the tongue. The stimulation of the hypoglossal nerve at night after surgical implantation of a device has recently been reported as non-inferior to PAP therapy (Heiser et al., 2023), and associated with improvements in patient reported outcomes (Braun et al., 2023). However, this treatment is indicated only for individuals with moderate to severe OSA and an AHI between 15 and 100/hr, and a BMI ≤ 40kg/m2, intolerant to PAP therapy and without concentric pattern of collapse of the upper airway as determined by sleep endoscopy (U.S. Food and Drug Administration, 2023). The latter is an expensive procedure, and the list of indicators means there are limited alternative treatment options for certain OSA patients (e.g., with an AHI <15/h).

In 1999, a novel, non-implantable therapy based on neuromuscular electrical stimulation (NMES) of the tongue’s motor nerves was developed as an alternative to positive airway pressure (PAP) treatment for patients with obstructive sleep apnea (Wiltfang et al., 1999). This approach primarily targets the genioglossus muscle, aiming to reduce the likelihood of upper airway collapse during sleep. A commercial device utilizing this mechanism, the eXciteOSA® (Signifier Medical Technologies) received FDA approval in 2021. A number of uncontrolled (Baptista et al., 2021; Kotecha et al., 2021; Wessolleck et al., 2018) and one randomized placebo-controlled trial (Abreu et al., 2023) provide support for this alternative treatment option for patients with mild to moderate OSA, in terms of improving snoring, sleepiness, sleep quality and AHI severity. The present study aims to add to this emerging literature by examining the impact of treatment on sleep health and functional outcomes in real-world patients given the long-standing evidence of impairment (Garbarino et al., 2020; Walia et al., 2017) with OSA established as an independent risk factor (Kim et al., 2019).

Specifically, the following questions were addressed: Q1: is there a pre-to-post change in patient reported outcomes (PROs). Q2: Is there a dose-response relationship between the treatment variables (adherence, intensity and intensity variance) and outcome (patient reported outcomes). We hypothesized an improvement in sleep disturbance, a reduction in sleep-related impairment and an improvement in depressive symptoms, after treatment with the device, and a dose-response relationship to be present between treatment variables and patient outcomes.

## Methods

### Study Design Overview

The study design is a retrospective observational study using real-time data collected through the eXciteOSA® app. Data were recorded prospectively by users during routine use of the app and were accessed for this analysis under an ethics approval by the Department of Psychological Sciences and Health ethics committee (79.19.07.2023.A). Data on patient reported outcomes, attention/vigilance, treatment related factors and demographics were extracted.

### Selection of participant data

Participants who provided at baseline and at least one post-treatment questionnaire were selected. Any follow-up data provided between 30^th^ of Jan 2022 and the 30th of April 2024 were included in the dataset, data extraction occurred on the 1^st^ of May 2025. No other inclusion criteria were set at the data extraction stage. At the time of data extraction, there were 14,443 eXciteOSA® users.

Individuals who did not complete the 42 sessions or did not have either a baseline and post-treatment questionnaire were excluded from the analysis (n=12,948). There was a wide range in the duration between the date of post-treatment questionnaire completion and finishing the 42 sessions.

Consequently, and to gather a “valid” post-treatment assessment of PROs, we included in the analysis only those individuals who completed a post-treatment questionnaire within 1 month of finishing their 42 sessions. This approach left us with a final sample of 469 individuals, see Figure 1 for details.

**Figure 1.**
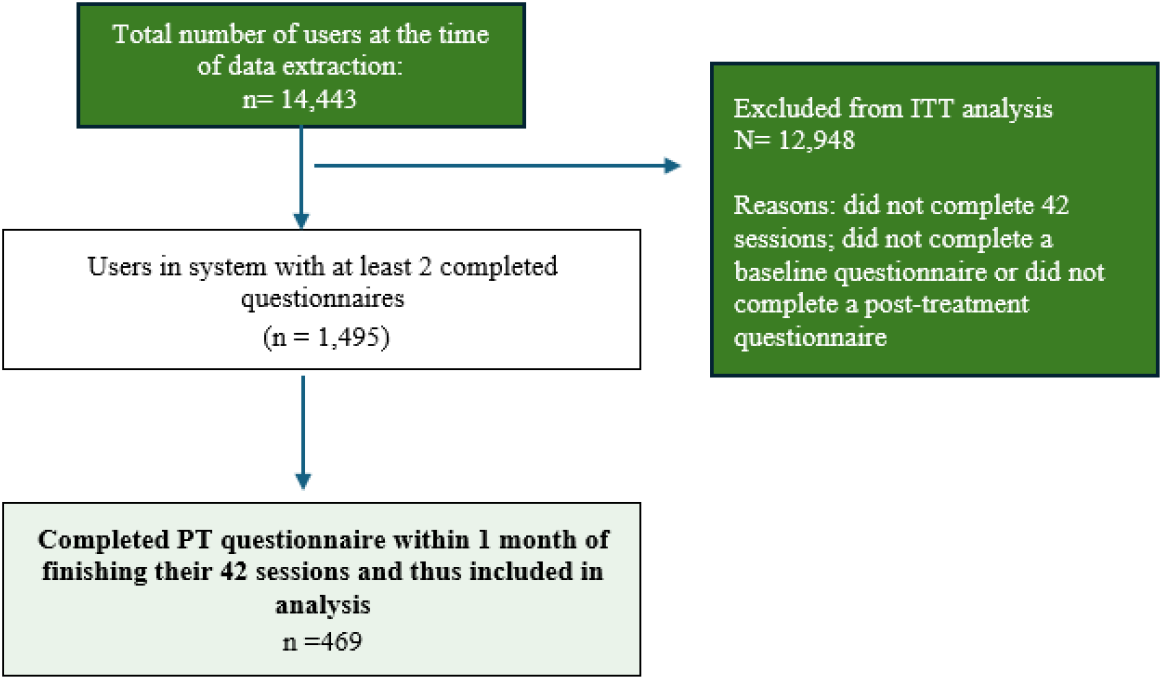
Consort Flow Chart

### Measures

PRO measures included the following:

The Functional Outcomes of Sleep Questionnaire 10 is the short version of the full scale. Through ten items it assesses impact of sleepiness on daytime functioning (Chasens et al., 2009; Weaver et al., 1997), such as “Do you have difficulty concentrating on the things you do because you are sleepy or tired?”. scores range from 1 (extreme difficulty) to 4 (no difficulty) with higher scores indicating less impairment. For some items, individuals can indicate they do not engage in a particular activity. The Patient Health Questionnaire 8 item version (PHQ-8) assesses mood without the suicidal thoughts question included in the full scale (Kroenke et al., 2001; Kroenke et al., 2009). It includes items such as “How often during the past 2 weeks were you bothered by little interest or pleasure in doing things?” and items are rated on a scale from 0 (not at all) to 3 (nearly every day), higher scores thus indicating worse mood. Two Patient Reported Outcome Measures Information System (PROMIS) measures were used: Sleep Disturbance to assess sleep disturbances (“In the past 7 days, I had a problem with my sleep”) and PROMIS-Sleep Related Impairment to assess daytime impact of sleep problems (e.g., “In the past 7 days, I had a hard time getting things done because I was sleepy”). The 4-item version of each scale was used and items included and scores ranged from 1 to 5 with higher scores indicating worse performance. Raw scores were converted into a T-score following the PROMIS scoring manual and conversion tables, there were no missing items (PROMIS Health Organization, 2023). Finally, the Work Productivity and Activity Impairment questionnaire(Reilly et al., 1993) was included to assess impairment in work productivity and activity with items such as “During the past seven days, how many hours did you miss from work because of your health problems?”, scoring followed guidelines (Reilly Associates, n.d.), of the subscales only overall work productivity is reported in the results. In addition to these PRO measures, information on demographics and treatment-specific information was extracted, see table 1 for details. PVT was included in the data collection, but not analyzed, see explanation in results for details.

**Table 1:** Overview of variables measured.

| **VARIABLES** | BASELINE | POST-TREATMENT | ANY SUBSEQUENT FU |
|---|---|---|---|
| DEMOGRAPHICS |  |  |  |
| Age | X |  |  |
| Gender | X |  |  |
| Ethnicity | X |  |  |
| Country | X |  |  |
| Weight & Height | X |  |  |
| TREATMENT-RELATED DATA |  |  |  |
| Average intensity |  | X | X |
| Intensity range |  | X | X |
| PRO MEASURES |  |  |  |
| Functioning (FOSQ-10) | X | X | X |
| Sleep disturbance (PROMIS- SD) | X | X | X |
| Sleep-related impairment (PROMIS-SRI) | X | X | X |
| Work performance (WPAI) | X | X | X |
| Depression (PHQ8) | X | X | X |
| PERFORMANCE |  |  |  |
| Psychomotor vigilance task (PVT) | X | X | X |

### Data storage

The data were extracted by Signifier Medical Technologies, and the encrypted and password-protected file was shared with the two investigators involved in the analysis (AJ and MC) safely through Strathclyde One Drive.

### Treatment

The device (eXciteOSA®, Signifier Medical Technologies) delivers intraoral stimulation for 20 minutes daily over an initial 6-week period, followed by twice-weekly sessions to sustain therapeutic benefits. The electrical stimulation is applied during the day to enhance tongue muscle tone and endurance, with residual effects that help maintain airway patency at night. The patient starts each 20-minute session by controlling the device through an app. Before the first session (and up to the start of the third session), the patient is asked to complete a brief survey about their sleep, mood and daytime functioning. This same survey is completed at the end of treatment (after 42 session) and at regular intervals thereafter (~90 days).

### Data preparation and analysis

For the PRO measures a total score was created for the FOSQ-10, the PHQ8 and the WPAI. For the PROMIS (sleep disturbance and sleep related impairment), a T score was computed. For the PVT, we were unable to compute a viable PVT score for all users, as access to the raw data was not possible through the app. This approach prevented trial-level data to be extracted that would allow for data cleaning and calculation of variables (mean reciprocal) that is expected as part of PVT data collection and reporting (Basner & Dinges 2011). To answer the two research questions (is there a pre-to-post change in patient reported outcomes (PROs) and is there a dose-response relationship between the treatment variables (adherence, intensity and intensity variance) and outcome (patient reported outcomes), a Bayesian linear mixed model was applied. The models included treatment adherence, treatment intensity and intensity variance, age and gender as co-variates, and tested the interaction between pre-and-post timepoints and each PRO questionnaire. As our model is Bayesian, we were able to compute the posterior probability of effects being in a given direction, rather than the probability of an effect if there were no difference, as in null hypothesis significance testing. For example, we can compute the probability that post-treatment scores are on average greater than pre-treatment scores for our PRO variables. We also explored how many individuals report symptoms below clinically relevant cut-offs at post-treatment. To answer this question, we applied posterior-predictive simulation, a Bayesian statistical approach that generates many new datasets from a fitted model. These datasets represent distributions of individuals, rather than means, and allow for the estimation of the probability that an individual starting above a particular cut-off would fall below it after treatment. For the two PROMIS scales (sleep-related impairment and sleep disturbance), the cut-off score was 55, for the PHQ-8 a cut off of ≥ 10 was considered for current depression (Kroenke et al., 2009) and for the FOSQ-10, a cut off of 17.9 was considered, with lower scores indicating worse functioning (Weaver et al., 2007). This approach allowed us to test the influence of various attributes on changes in PRO scores and return probabilities for new participants with the characteristics of interest of completing treatment below the cut-off. Each of the PRO questionnaires was subjected to factor analysis and parallel analysis to check their psychometric properties against their initially reported structures, with each questionnaire showing the same factor structures as their initially published estimates.

## Results

### Descriptive statistics

Individuals included in the analysis were 56 years old (SD 13.6), predominately male (59%), resident in the USA (35%), white (85%) and with a BMI of 29.1 kg/m2 (SD 8.6). Table 2 outlines the baseline and post-treatment scores for sleep disturbance, sleep-related impairment, depression, work impairment, and daytime functioning.

**Table 2:** Model-implied Baseline and Post-treatment Scores.

|  | Baseline | Post-treatment | Mean difference | 94% HDI |
|---|---|---|---|---|
| PROMIS-SD | 55.3 (0.65) | 49.10 (0.67) | -6.2 | -8.44 to -3.86 |
| PROMIS-SRI | 52.70 (1.28) | 42.57 (1.31) | -10.12 | -14.51 to -5.54 |
| PHQ8 | 7.30 (0.50) | 3.79 (0.51) | -3.51 | -5.35 to -1.87 |
| FOSQ-10 | 16.22 (0.30) | 16.35 (0.30) | 0.13 | -0.91 to 1.17 |
| WPAI - Absenteeism | 6.22 (1.39) | 1.15 (1.42) | -5.07 | -9.69 to –0.09 |
| WPAI - Impairment | 29.74 (2.58) | 15.97 (2.66) | -13.77 | -22.81 to –4.86 |
| WPAI - Presenteeism | 24.79 (2.39) | 11.79 (2.47) | -13.00 | -21.23 to –4.76 |
| WPAI – Work impairment | 25.98 (2.58) | 12.75 (2.66) | -13.23 | -21.73 to –4.04 |

*Note*. HDI = highest density interval; PROMIS-SD = Patient-Reported Outcomes Measurement Sleep Disturbance; PROMIS-SRI = PROMIS Sleep-Related Impairment; PHQ-8 = Patient Health Questionnaire-8; FOSQ-10 = Functional Outcomes of Sleep Questionnaire-10; WPAI = Work Productivity and Activity Impairment.

### Improvement in PROs at posttreatment

Comparisons of average scores on the patient-reported outcome (PRO) measures revealed credible improvements in depression (100% chance there will be an improvement at post-treatment), sleep disturbance (100% chance), sleep-related impairment (100% chance), presenteeism (99.8% chance), activities impairment (99.7% chance). There was no credible improvement at post-treatment in functional outcomes related to sleepiness (60.9% chance of improvement). These changes were examined by controlling for time between the first and last questionnaire completion, adherence, treatment intensity, and variability in treatment intensity as well as age and gender. The average difference scores between baseline and post-treatment and highest density intervals (HDI) are listed in Table 2 and the visual representation of this difference for each measure is included in Figure 2.

**Figure 2.**
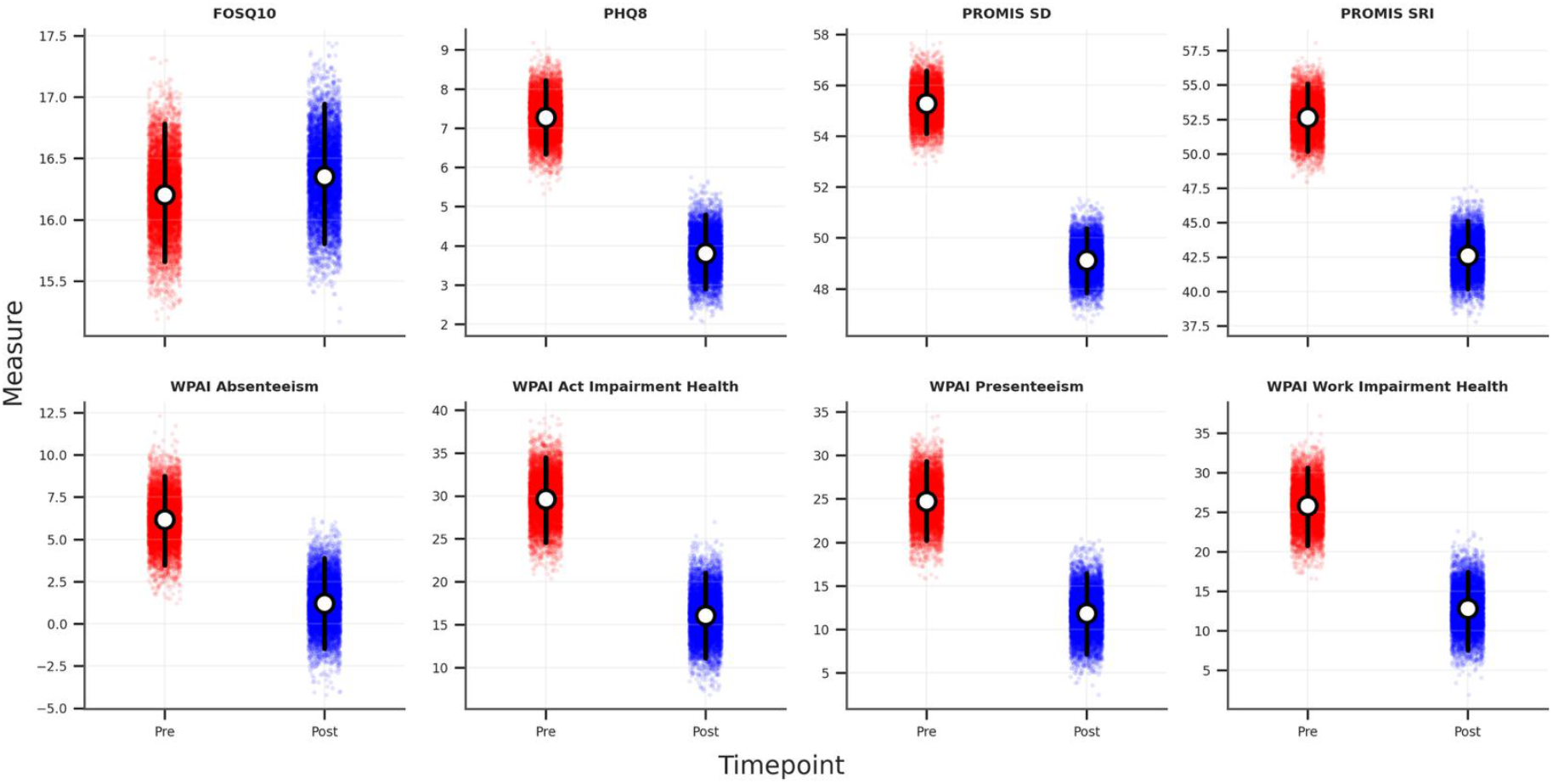
Pre to Post changes in patient reported outcomes Each quadrant includes the baseline (red), and post-treatment (blue) predicted means (white dot) along with the credible intervals (black bar) for these means. The red and blue dots are credible means for each variable at each time point.

We also examined whether there was a dose–response relationship between adherence (dose) and PRO outcomes (treatment response), figure S1-4. This relationship only appeared to be the case for functional impairment related to sleepiness (FOSQ) and variance in treatment intensity and adherence, see figure S1, S3 and S4 in the supplemental file where the 95% confidence intervals excluded zero. This effect is in the predicted direction (more treatment dose/less intensity variability was associated with more improvement); however, given the absence of a pre-to-post treatment effect for this variable (see Figure 2), the finding maybe spurious and should not be interpreted as evidence of a meaningful dose-response relationship.

Lastly, we were also interested in clinically meaningful improvement by estimating the proportion of individuals whose post-treatment scores fell below validated clinical cut-offs. Using posterior predictive simulations derived from the Bayesian hierarchical model presented above, we estimated the probability that an individual with a given baseline score would achieve a post-treatment score indicative of remission (i.e., below the impairment threshold). For each outcome, we generated posterior predictive distributions of post-treatment scores conditional on fixed baseline values (these fixed values were just above [or below for the FOSQ] the published cut off scores for impairment) and computed the proportion of simulated observations below (or above for the FOSQ) the corresponding clinical cut off. Among simulated sample of individuals with a baseline score of moderate sleep disturbance and moderate sleep-related impairment (band of 60-62), the posterior probability of a post-treatment score below 55 (normal limits) was 86% and 87% respectively. For depression, the probability of a clinically meaningful improvement, defined here as a baseline score of 11-13 and a post-treatment score <10 was 91%. For functional outcomes related to sleep, the posterior probability of having a baseline score between 16-17 and ending with a score above 17.9 (for the FOSQ, higher scores indicate better functioning) was 26%, see Figure 3.

**Figure 3.**
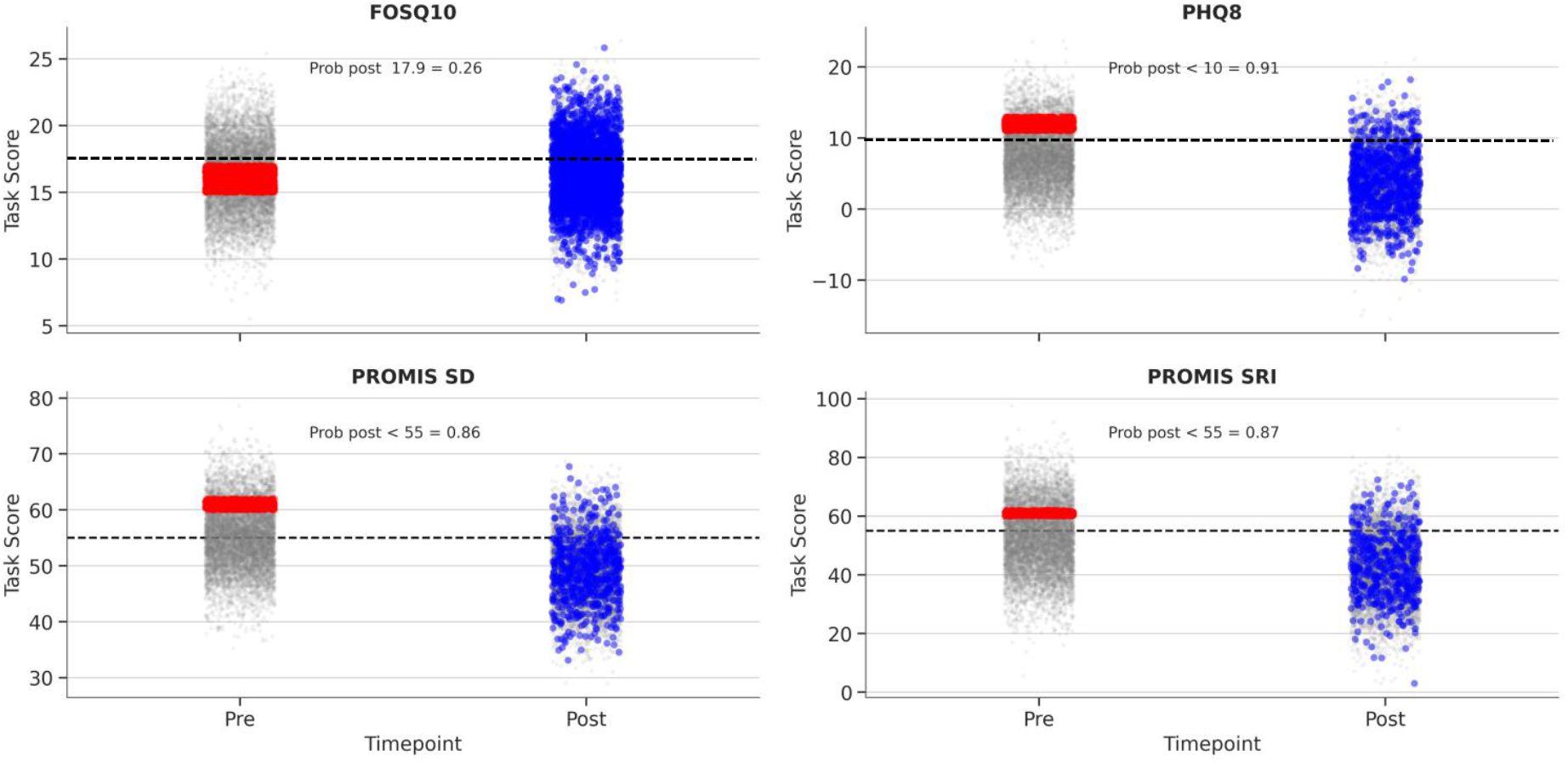
Posterior-predictive simulation depicting likelihood of scoring below (or above for FOSQ) cut off scores at post-treatment. Red dots indicate individuals with a pre-treatment score within a selected range. Blue dots indicate the post-treatment score of all these individuals. Probability indicates the likelihood that these blue dots (scores) are below the published cut off.

### Exploratory Analysis

Because prevalence rates of sleep disorders such as insomnia (Endomba et al., 2023) or OSA (Ghavami et al., 2023) and depression (Zhang et al., 2024) are higher in older adults, we conducted exploratory analysis to understand the effects in those ≥ 65 years. Predicted scores for participants ≥65 years based on full-sample regression model were computed (131 participants in the full sample were ≥65 years). Results remain largely unchanged, although the magnitude of impairment (both pre and post intervention) is slightly less marked than in the previous models, see figure S5 in the supplementary material.

## Discussion

The aim of this study was to examine the effects of neuromuscular electrical stimulation device on patient-reported outcomes in a real-world sample. Robust changes in depression, sleep-related impairment, sleep disturbance, and work impairment were evident, even after controlling for age, gender, adherence and treatment intensity (absolute and variance). Improvements in functional impairment related to sleepiness were not evident. This finding is likely due to the sample selection, (i.e. patients’ presenting with mild OSA or primary snoring) and is consistent with some of the other clinical trials evaluating non-PAP therapies with the FOSQ as an outcome (de Ruiter et al., 2018; Marklund et al., 2015). The improvements in total scores and the percentage of individuals meeting criteria for diagnosis versus within “normal” limits are encouraging and supplement findings from smaller, clinical research samples (Baptista et al., 2021; Kotecha et al., 2021). The lack of a dose response relationship should be interpreted in the context that adherence in this sample was very good. Eighty-six percent of the sample completed their 42 sessions within 50 days of beginning treatment. Average intensity was 8.58, which is above what was considered therapeutic in the placebo control study (Abreu et al., 2023). Lack of variance in the “dose of treatment” limits the ability to understand fully these dose-response relationships. Relative to studies of PAP therapy, improvements in sleep-related symptoms were larger here than those reported in the only PAP study to use PROMIS measures ((Donovan et al., 2020) for both sleep disturbance (−6.2 vs −4.0 T-score points) and sleep-related impairment (−10.1 vs −4.2). The reduction in depressive symptoms (PHQ-8: −3.5 points) was smaller than in PAP cohorts using the PHQ-9 (mean −7.6 in (Edwards et al., 2015); median 10 to 5 at six months in (Velescu et al., 2022)). In contrast, functional outcomes showed little change (FOSQ-10: +0.13); smaller than gains reported following PAP in uncontrolled (+3.3

(Billings et al., 2014) and randomised controlled studies (+0.98 (Weaver et al., 2012). To our knowledge, no studies have reported WPAI outcomes following PAP therapy. These comparisons should be interpreted with caution as PAP therapy is a different treatment, the comparator samples included only OSA patients (not a mixture of OSA and primary snorers as this study had) and consequently had higher baseline symptom levels, leaving more room for improvement; and different questionnaire versions were used, all of which could account for observed differences.

The study results should be interpreted in light of both strengths and limitations. The sample size and real-world data were strengths. The sample size was larger than any previous study published evaluating this device. The real-world nature captures data from individuals who are not selectively agreeing to participate in clinical trials, which usually includes more educated, female, and less symptomatic individuals who have also been evaluated against stringent inclusion/exclusion criteria. Thus, this real-world sample represents a broader clinical and perhaps more generalizable population. Lastly, validated questionnaires were used, including those where standardized scores are available to compare to normative values (e.g., PROMIS).

The limitations of this study included sample selection and lack of control group. We were only able to analyze data from individuals who had completed at least two questionnaires on the app. While the baseline question was compulsory, this requirement was not the case for the VA users (approx. 20% of all users). The need to complete questionnaires was removed for all users referred through the VA system in the USA. We also did not have data on OSA severity or which of the participants had mild OSA vs primary snoring, which could have been an important covariate. Without a placebo control group, we were unable to rule out effects such as regression to the mean or expectation of change.

To estimate the contribution of regression to the mean (RTM) to the observed improvement in depressive symptoms, we adapted Linden’s (2013) equations 5 and 6. In Linden’s approach, the distance between a subgroup’s baseline mean and the population mean (Cσ) is estimated from the cut-off used to select that subgroup; as participants in the present study were not selected on PHQ-8 score, this distance could instead be calculated directly from the observed baseline mean (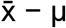).

Substituting this value into Linden’s equation 6 gives an expected post-test mean in the absence of treatment of μ + ρ(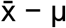), with the expected RTM effect being the difference between this value and the observed baseline mean. We used a reference mean (μ) of 6.6, the baseline mean of a large sample of adults with chronic conditions (Ory et al., 2013), and an 11-day test-retest reliability (ρ) of .83 for the PHQ-8 total score (Mattsson et al., 2020). With the pre-test mean of 7.3 and a post-test mean of 3.8, this calculation means the estimated regression to the mean is 0.12, which is less than 4% of the observed change of 3.5. The improvement in depressive symptoms is therefore unlikely to be explained by RTM. The same approach was applied to the PROMIS scales, using the general population reference mean of 50 on the T-score metric (PROMIS Health Organization, 2023). To our knowledge no test-retest reliability values for the 4a short forms are available; thus, we used values from other short forms from an untreated control arm of a clinical trial on TENS for women with fibromyalgia (PROMIS Sleep Disturbance short form 8b, ρ = .71; PROMIS Sleep-Related Impairment short form 8a, ρ = .62; (Chimenti et al., 2021)). The expected change attributable to RTM was 1.5 points for Sleep Disturbance (25% of the observed reduction of 6.2 points) and 1.0 point for Sleep-Related Impairment (10% of the observed reduction of 10.0 points). Moreover, post-treatment means on both scales fell below the population mean of 50, a change that RTM alone cannot produce. RTM may therefore have contributed modestly to the improvement in sleep disturbance but is unlikely to account for the improvements observed on either PROMIS scale. For the WPAI, RTM was estimated for overall work impairment only (for simplicity), using the mean of non-OSA controls in the 2016 US National Health and Wellness Survey as the reference (16.9%; (Stepnowsky et al., 2019)) and a 3-month test-retest reliability from a placebo-control arm with individuals with migraine (ICC = .36; (Ford et al., 2023)Estimating the effects from expectation of change could also be reliably captured with a future randomised placebo-controlled trial.

Despite these limitations, this report provides evidence for the impact of a neuromuscular electrical stimulation on patient-reported outcomes. The findings demonstrate that NMES therapy is associated with clinically relevant improvements in multiple patient-reported outcomes in a large real-world cohort. These results add to the evidence base established by prior randomised and observational studies supporting NMES as a therapeutic option for individuals with mild OSA or primary snoring.

The results need to be supplemented with future placebo-controlled trials using both subjective and objective measures of functioning.

## Data Availability

The data used in this study were obtained from Signifier Medical Technologies and are proprietary; they are not publicly available and cannot be shared by the authors. Requests for access to the data should be directed to Signifier Medical Technologies, subject to their approval and data sharing agreements.

## CRediT statement

Conceptualization: MC, AJ, CO

Data curation: AJ, CO, MC

Formal analysis: AJ

Funding acquisition: N/A

Methodology: MC, AJ, CO

Project administration: MC

Resources: CO

Visualization: AJ

Writing – original draft: MC, AJ, CO, SP, AM, HW, JO

Writing – review and editing: MC, AJ, CO, SP, AM, HW, JO

## Conflict of interest statement

MC and AJ received consulting fees from Signifier Medical Technologies for the analysis and preparation of the manuscript. Analysis was conducted independent of Signifier. AM reports income from the NIH. He has consulting income from Eli Lilly, LivaNova, Zoll, Aerin and Sunrise. He is co-founder and has equity in Clairyon, a small startup related to predictive analytics. Resmed gave a philanthropic donation to UCSD. HW served on advisory board of Resmed, Eli Lily, LivaNova. SP reports grants from NIH and PCORI. He has consulting income from Fisher & Paykel, Zoll-Respircardia, Inc., Abbvie, Inc., Inspire Medical Inc., and Jazz Pharmaceuticals, Inc. in the past 36 months.

## Notes

### Author Declarations

Department of Psychological Sciences and Health ethics committee from the University of Strathclyde gave ethical approval for this work (79.19.07.2023.A).

